# Understanding Community Knowledge and Practices for *Taenia solium* Taeniasis and Cysticercosis: Baseline findings for Pig farmer communities of Punjab, India

**DOI:** 10.1101/2025.06.01.25328727

**Authors:** Rashmi Sharma, Rajnish Sharma, Rabinder Singh Aulakh

**Author notes:** Corresponding author-Dr. R.S. Aulakh, Professor and Director Student Welfare (DSW), Guru Angad Dev Veterinary and Animal Sciences University, Ludhiana, Punjab, 141004, Email id.

## Abstract

**Objective:** The study was aimed to assess knowledge, attitude and practices (KAP) regarding *Taenia solium* taeniasis and cysticercosis among pig farming communities in Punjab, India

**Methods:** Cross-sectional survey of 719 pig farmers across five districts was conducted using a structured questionnaire comprising of 35 questions to assess the knowledge, attitude, and practices. Univariate logistic regression was performed to identify factors associated with good knowledge (≥30).

**Results:** Overall, knowledge was critically low (mean 14.32 ±8.51). only 2.8% achieved good knowledge, 83.30% heard of taeniasis versus 5.70% of cysticercosis. Pig regarding practices significantly predicted good knowledge (p=0.010), with intensive systems showing 3.46 times higher odds than free-range systems (OR=3.464, 95% CI0.979-12.261). Occupation approached significance (p=0.54). despite 100% reporting handwashing, knowledge-practice correlation was non-significant (r=0.033, p=0.378). Only 8.8% willing to participate in mass drug administration programmes.

**Conclusion:** Critical knowledge gaps regarding *T. solium* transmission persist in Punjab’s pig farming communities, Targeted visual-based health education for low-literacy population and infrastructure improvement are urgently needed.

## Introduction

*Taenia solium taeniasis* and cysticercosis (TSTC) has a significant global public health and economic burden, particularly in low-income and middle-income countries (LMICs) (Mkupasi *et al*., 2024; Franco-Muñoz C *et al*., 2025). *T. solium* is ranked as the foodborne parasite of greatest concern worldwide by the FAO and WHO, TSTC impacts approximately 50 million people annually (Nyangi *et al.*, 2024; Franco-Muñoz C *et al*., 2025). The burden is primarily driven by the neurological consequences of the parasite’s larval stage, which accounts for nearly 30% of acquired epilepsy cases in endemic regions across sub-saharan Africa, Asia and Latin America (Millogo *et al*., 2019; Onzivua *et al*., 2026; Marwa *et al*., 2023).

The parasite follows a complex indirect life cycle involving humans as the sole definitive hosts and pigs as the primary intermediate host (Nyangi *et al*., 2024; Franco Muñoz C *et al*., 2925). Human acquire taeniasis by consuming raw or undercooked pork containing viable cysticerci, which mature into adult tapeworm in the small intestine (Nyangi *et al*., 2024). These worms release gravid proglottids containing thousands of microscopic eggs into the environment via human faeces (Marwa *et al*., 2023). Pigs become infected with porcine cysticercosis by ingesting these eggs from contaminated soil or waste (González *et al*., 2003). Accidental human ingestion of eggs through the fecal-oral route or autoinfection leads to human cysticercosis. If larvae migrate to the central nervous system, they cause neurocysticercosis (NCC), the leading preventive cause of epilepsy in endemic regions (Nyangi *et al*., 2022; Lugelongi *et al*., 2025; Figure-1).

Despite being designated as potentially eradicable, TSTC persists due toa complex interplay of environmental, behavioural, and infrastructural risk factors (Bibi *et al*., 2023). These include inadequate sanitation, free-range pig management, and insufficient meat inspection (Wilson *et al*., 2023). Crucially, the maintenance of the infection cycle is driven by significant knowledge gaps regarding transmission (Nyangi *et al*., 2025; Makingi *et al*., 2023), Studies across regions such as Pakistan, Ethiopia and Tanzania indicate that while communities may recognise “measly pork”, they often lack a scientific understanding of the disease spreads (Bibi *et al*., 2023). In the Asian region, Knowledge, attitude and practice (KAP) studies have proven invaluable for identifying these local socio-epidemiological drivers and identifying high-priority demographics for interventions (Nanda *et al*., 2014; Mishra *et al*., 2007). Deeply ingrained cultural myth further complicate control, for instance, in north-west India that cabbage consumption causes cysticercosis often overrides scientific evidence (Nanda *et al*., 2014; Mishra *et al*., 2007)

The primary objective of this study was to evaluate the level of knowledge, attitude, and practices regarding *T. solium* taeniasis and cysticercosis among pig-farming communities across five districts of Punjab. The mapping of awareness gaps and identifying risk husbandry and dietary behaviours, this research provides the necessary evidence base to design community-centred educational packages. These finding are intended to guide public health policy toward multi-sectoral elimination strategies tailored to the local cultural and occupational context.

## Methods

### 2.1 Study design and population

It was a cross-sectional study to evaluate pig farmers knowledge, attitude, and practices *T. solium* cysticercosis taeniasis and cysticercosis using a structured questionnaire. The study was conducted between July, 2019 to January, 2020 and included 750 participants (assuming that 50% of population possess the factor of interest with 5% absolute precision and 95% confidence interval, Dhand *et al*., 2014; Statulator) in the state of Punjab, India. A multistage cluster sampling methodology was implemented to recruit study participants. First, 5 districts were randomly selected. Subsequently, 5 taluka’s per district’s were randomly recruited. Finally, pig farming communities present within the selected taluka’s were purposively recruited. Communities were eligible for inclusion if atleast 10% of selected individuals of community are actively engaged in pig farming (Figure 2). Individuals primarily associated with pig farming were targeted for inclusion into the study.

**Figure 1:**
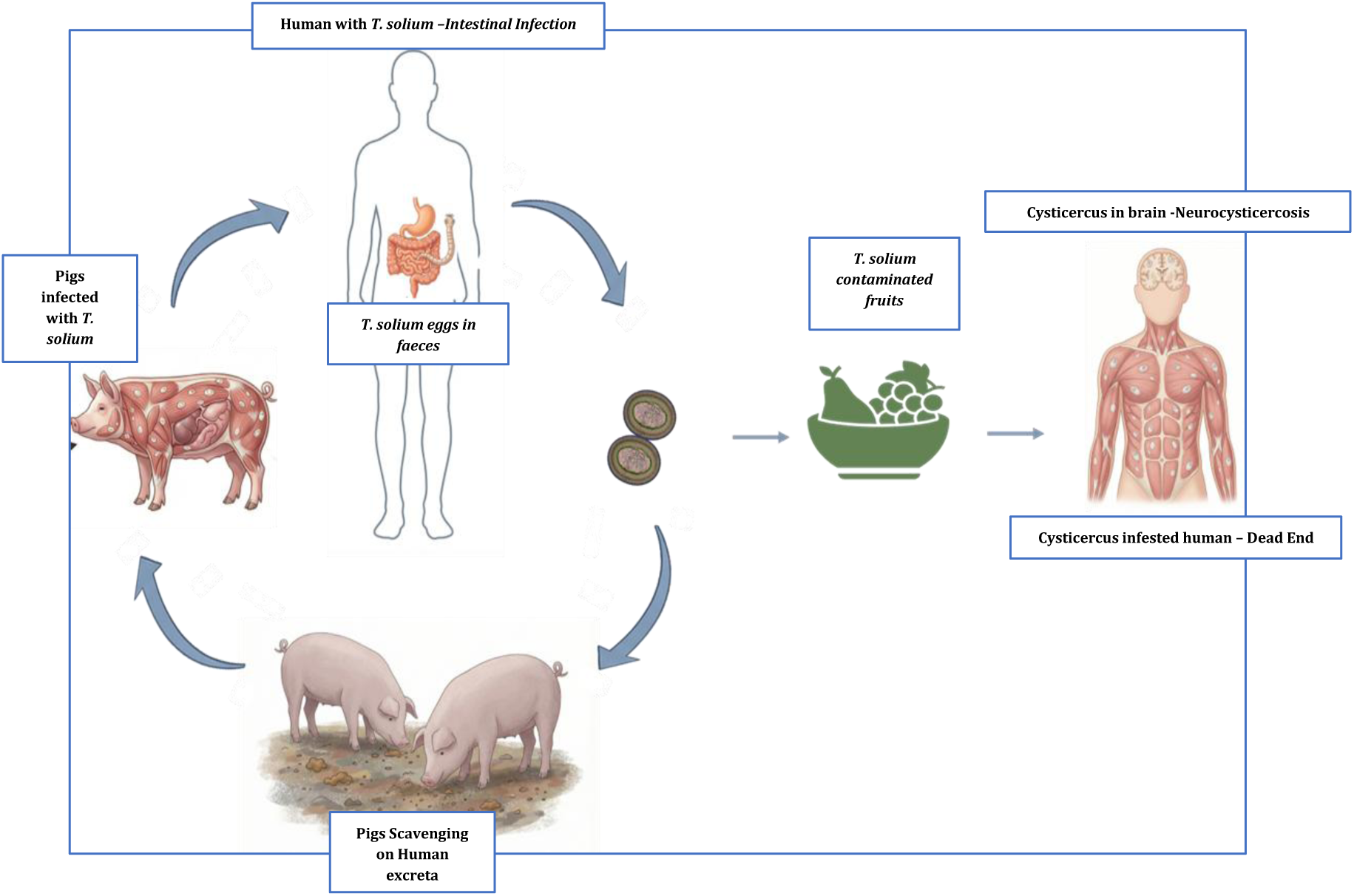
Lifecycle of *Taenia solium* Taeniasis and cysticercosis: Identifying the critical transmission points in Human-Swine interface

**Figure 2:**
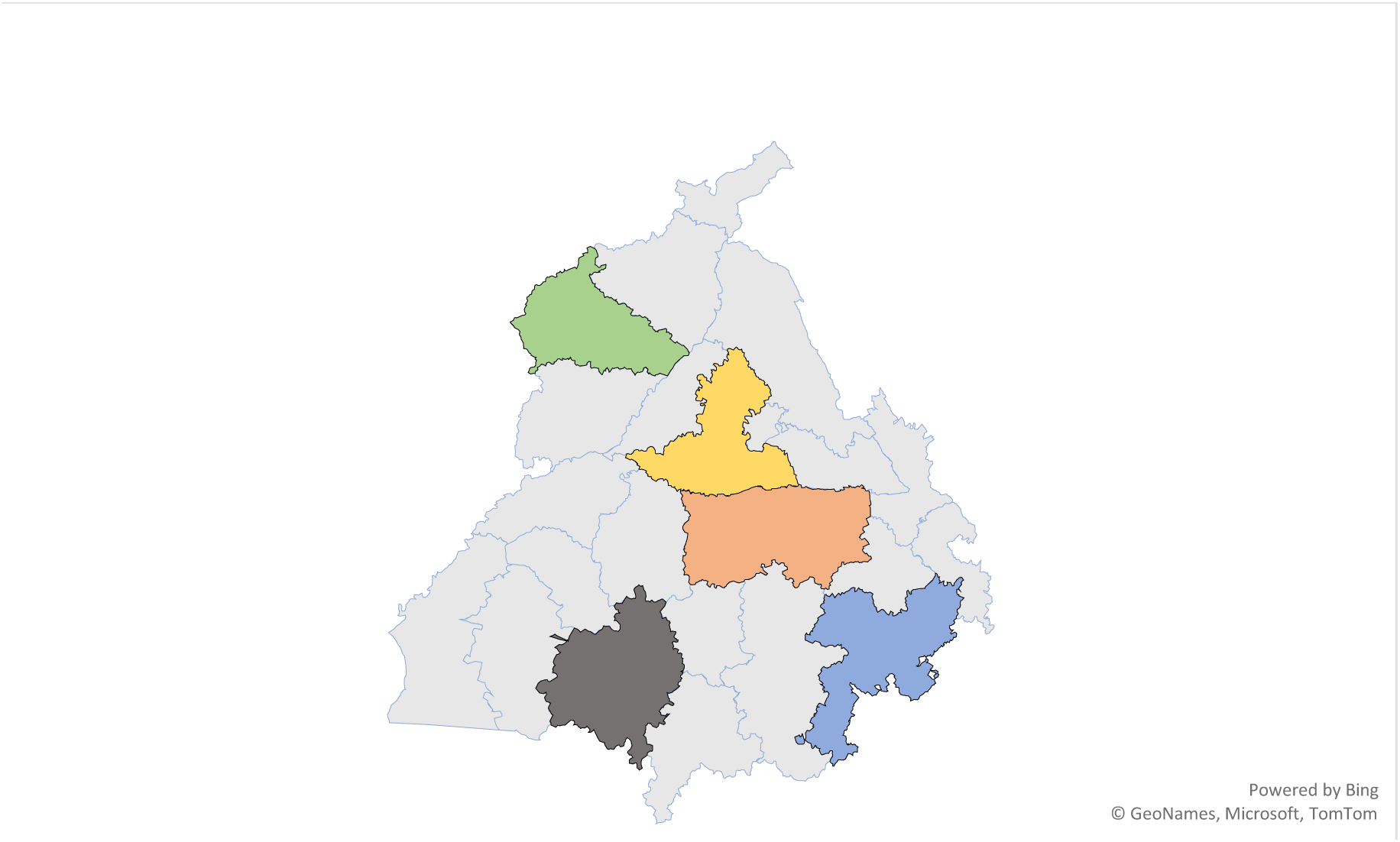
Geographic distribution of the selected study area for knowledge, attitude and practices study on *Taenia solium* taeniasis and cysticercosis (July, 2019-January, 2022)

### 2.2 Questionnaire

A written informed consent was obtained from all the participants before administration of questionnaire. To ensure linguistic and contextual appropriateness, survey terminology was adapted to align with local language and colloquialisms used in pig farming communities. This adaptation was validated through participatory consultation with community members, local health workers, and indigenous leaders. Data collection was conducted through face-to-face interviews among the selected participants. However, due to restrictions imposed by the COVID_19 pandemic, a subset of interviews (273) was conducted via telephone.

A comprehensive structured questionnaire was developed to assess respondents’ understanding of *T. solium* taeniasis and cysticercosis with particular emphasis on transmission routes, prevention practices, and animal husbandry. The questionnaire included both close-ended and open-ended questions. The final questionnaire consisted of 35 questions organised into three principle domains: Knowledge (15 questions), Attitude (8 questions) and Practices (12 questions). The Knowledge domain assessed general awareness of taeniasis and cysticercosis, understanding of their causative agents, and with particular emphasis transmission pathways, including items on how humans acquire tapeworm infection, the mode of transmission leading to cysticercosis, and how pigs acquire infection, in addition to items on clinical symptoms and preventive practices such as deworming and personal hygiene.

Additionally, socio-demographic information including age, gender, education level, and occupation was collected to enable stratified analysis. The survey instrument was developed in both English and punjabi to accommodate participant language preference.

Prior to field implementation, the questionnaire underwent content validation by subject matter experts (the primary research supervisor and a parasitology specialist with experience in zoonotic disease). Expert feedback was incorporated to refine question clarity, relevance, and appropriateness. The survey responses were subsequently coded and digitized

### 2.3 Data management and Statistical analysis

Data collected through the KAP survey were entered into Microsoft Excel (2019) spreadsheet and subsequently transferred to IBM statistical Package for Social Sciences (SPSS) version 26.0.0 for analysis. Data quality was ensured by comparing electronically entered records against original hard copy questionnaires to identify and correct transcription errors.

Preliminary descriptive analysis was conducted to characterize the study population and describe the distribution of knowledge, attitude, and practices responses. Categorical variables were summarized as frequencies and percentages. Response rates across districts were calculated to assess survey coverage. Knowledge, attitude and practice scores were calculated for each participant and categorized as follows: Low ≤ 33, Medium ⁓ 34-66 and High score ≥67. The number and percentages of respondents in each category were calculated and reported as frequencies and percentages.

To assess associations between KAP variables and sociodemographic characteristics (age, gender, education level, occupation, and pig farming system intensity), univariate analysis was performed using appropriate statistical tools. Multivariate logistic regression models were constructed to examine independent associations between sociodemographic (independent variables) and KAP outcomes (dependant variable), adjusting potential confounders. Statistical significance was established at a p-value <0.05.

### 2.4 Ethical considerations

The study protocol was approved by the Institutional Ethics Committee of Dayanand Medical College, Ludhiana, Punjab (IEC Ref No. 2020-553). All procedures were conducted in accordance with institutional and national ethical standards.

## 3.0 Results

### 3.1 Demographic Characteristics of the study population

The study surveyed 719 respondents across five districts, achieving a response rate of 73.14%. Participation was distributed among Ludhiana (38.50%), Jalandhar (25.65%), Amritsar (15.13%), Patiala (9.59%) and Bathinda (9.17%) (Table 1). The cohort was predominantly male (73.85%) and a majority (60%) were aged 31 years or older, reflecting the primary demographic active in local pig farming sector.

**Table 1:** Socio-demographic characteristics of study participants in pig farming communities, Punjab, India (n=719)

| S. No. | Variable | Category | n | % |
| --- | --- | --- | --- | --- |
| 1 | Location | Amritsar | 116 | 16.13% |
|  |  | Bathinda | 66 | 9.17% |
|  |  | Jalandhar | 191 | 26.65% |
|  |  | Ludhiana | 277 | 38.50% |
|  |  | Patiala | 69 | 9.59% |
| 2 | Gender | Male | 531 | 73.85% |
|  |  | Female | 188 | 26.14% |
| 3 | Age (years) | 18-30 | 287 | 39.90% |
|  |  | 31-40 | 235 | 32.70% |
|  |  | 41-50 | 123 | 17.10% |
|  |  | 51-60 | 55 | 7.60% |
|  |  | >61 | 19 | 2.60% |
| **4.** | **Educational Qualification** | Grade 1-5 | 426 | 59.20% |
|  |  | Grade 6-8 | 200 | 27.80% |
|  |  | Grade 9-10 | 81 | 11.30% |
|  |  | Grade 11-15 | 12 | 1.70% |
| **5** | **Involved in pig rearing** | Yes | 393 | 54.70% |
|  |  | No | 326 | 43.50% |
| **6** | **Deworming in last 12 months** | No | 649 | 90.30% |
|  |  | Yes | 70 | 9.70% |

*Survey conducted July, 2019-January, 20021, across five districts of Punjab, India*

Occupationally, over half of the respondents (54.7%) were actively involved in pig farming. The education qualification of surveyed population was notably low with 59.20% participants studied uptill 5^th^ grade. Furthermore, the baseline health practices revealed was lack of preventive care, as 90.30% of respondents had not undergone deworming in the 12 months prior to the survey (Table 1).

### 3.2 Knowledge regarding Taeniasis and cysticercosis

A substantial disparity in disease awareness was observed among the study participants, as 83.30% had heard of taeniasis compared to only 5.70% for cysticercosis (Figure 3). Overall knowledge scores were critically low with a mean knowledge score of 14.32 ± 8.51 (median 13.02, range 0.0-47.4). Only, 2.8% of the total surveyed population achieved an overall knowledge score classified as good (≥30). Transmission knowledge was limited with less than 2% of respondents correctly identifying established infection routes. Participants reported specific misconceptions regarding the aetiology of the disease, notably associating cysticercosis with the consumption of cabbage.

**Figure 3:**
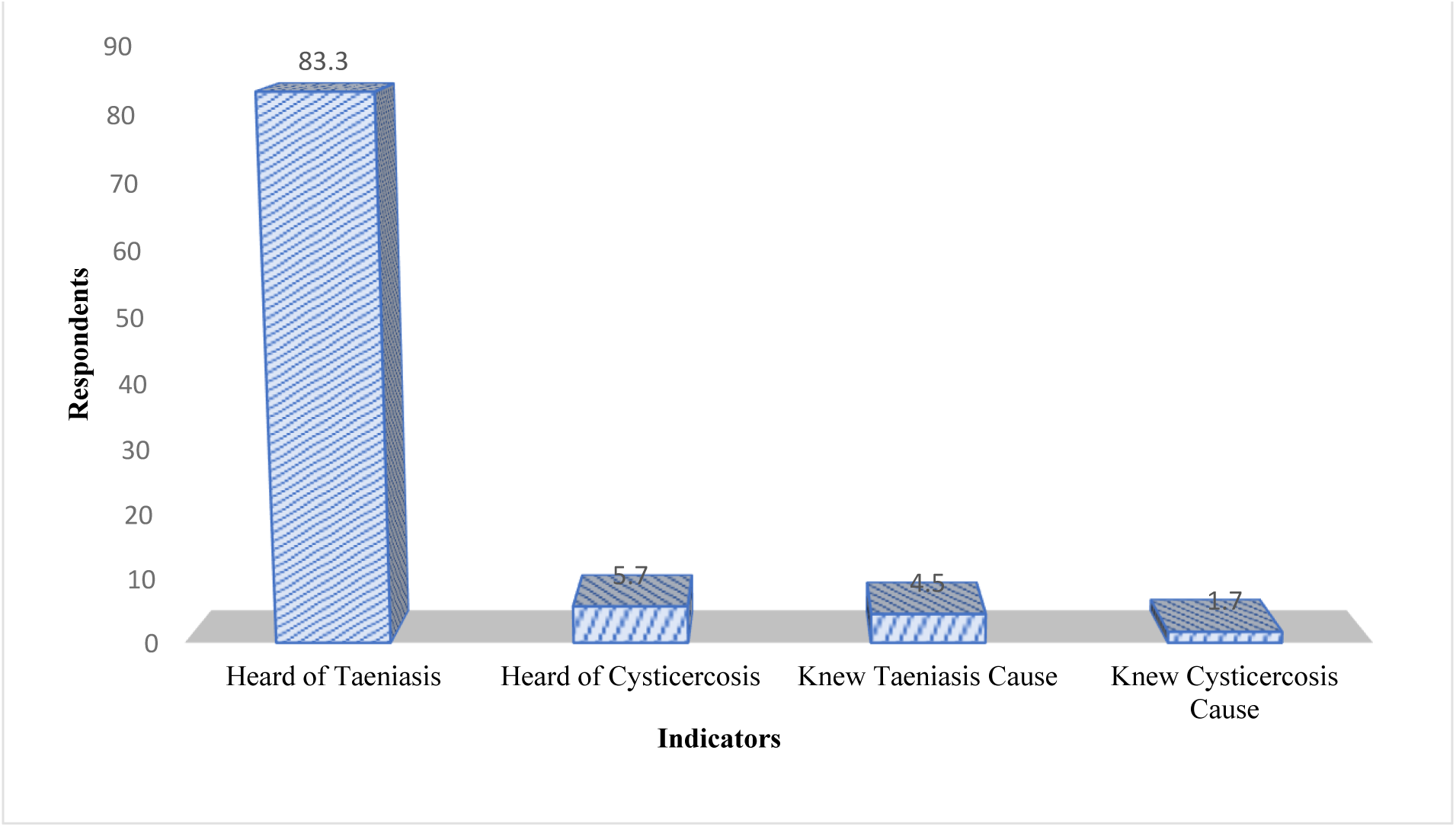
Illustrates the awareness disparity in *Taenia solium* infections in across key knowledge indicators among the surveyed population

Furthermore, 2% of the surveyed population does not characterize these conditions as disease, instead categorizing them as normal human physiology. Univariate logistic regression indicated that formal education level was not a predictor of awareness, as no participants with education above the matriculation level achieved a good knowledge score. Conversely, individuals involved in intensive pig rearing systems scored significantly higher than those using semi-intensive or extensive systems (Figure 4).

**Figure 4:**
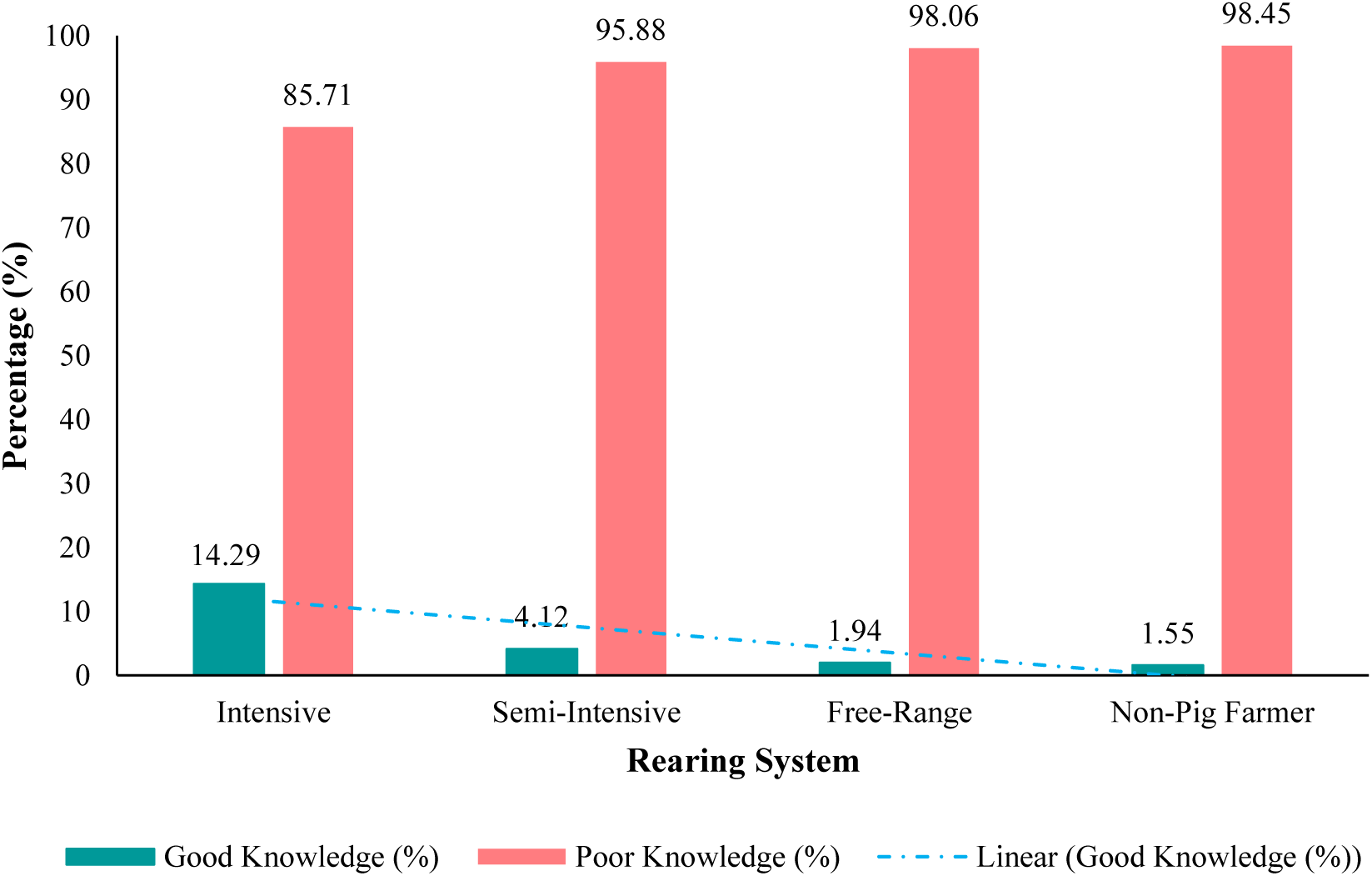
Demonstrates the association between pig management practices and knowledge scores across rearing systems

### 3.3 Attitude and Practice Towards Control and Management

Attitude scores demonstrated a mean of 39.45 ± 10.97 (median 37.50, range 12.5-75.00). While 67.2% of respondents characterized pig scavenging as a malpractice, 79.97% reported frequent observations of pig consuming garbage or litter in the vicinity of their residencies. The participant’s analysis of medical help seeking behaviour revealed that, in case of an 95.70% of participant’s would seek professional medical consultation with 80% preferring allopathic medical intervention over homeopathic/ayurvedic.

Although, 25.50% participants believed mass drug administration (MDA) could facilitate disease management, only 8.80% expressed a personal willingness to participate in such programmes, indicating a mass discrepancy in ideology and practice (Figure 5). Further, risk perception regarding meat safety was limited, as only 11.40% of respondents recognised the consumption of measly pork as a health hazard. While all respondents advocated for the necessity of household toilets, only 5.42% were aware of direct role played by human defecation in the transmission and persistence of infection cycle.

**Figure 5:**
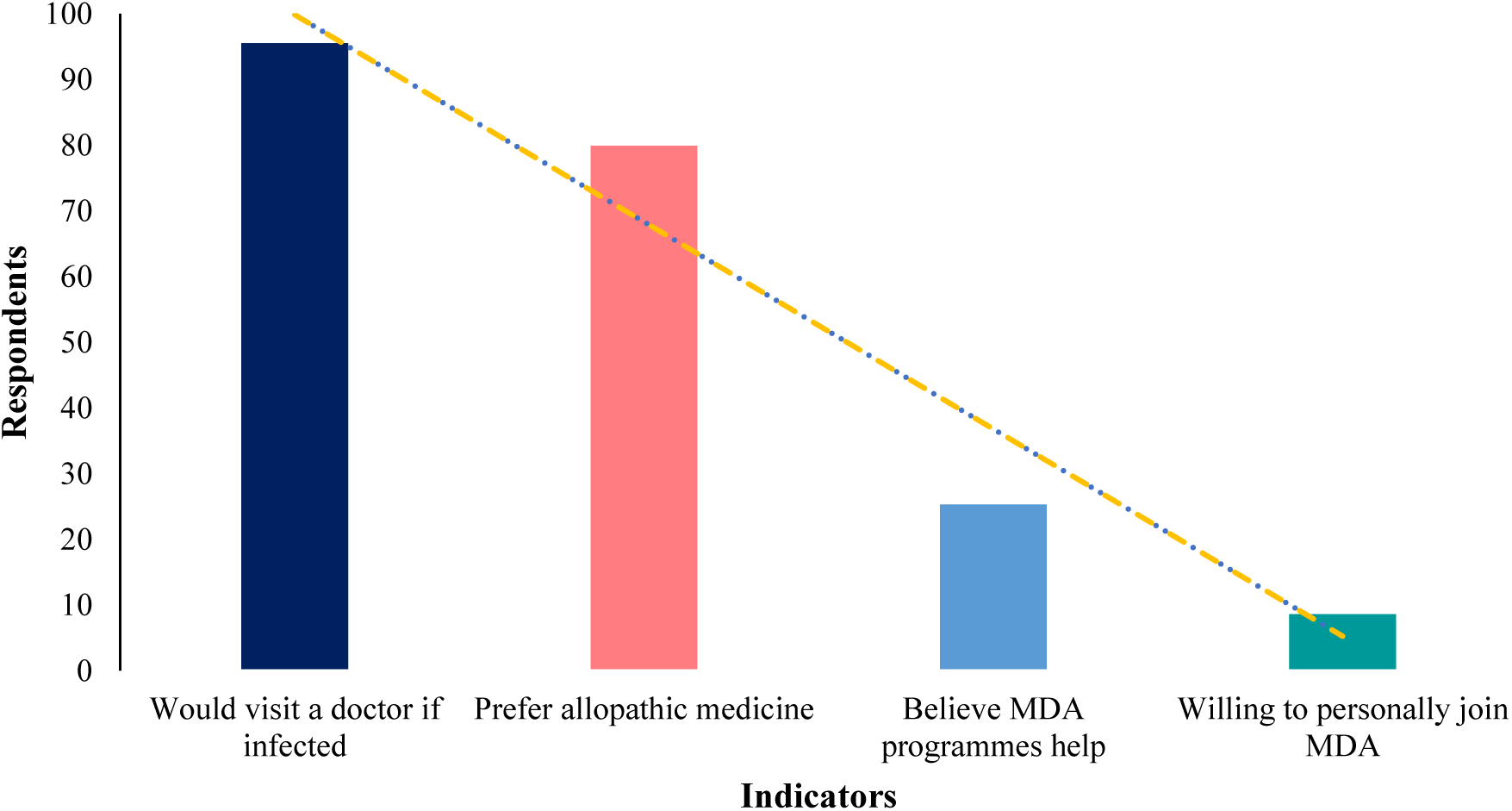
Highlight the variation between individual treatment seeking behaviour and collective participation in preventive public health programmes

The practice scores were higher than knowledge scores, with mean practice scores of 74.11 ± 15.51 (median 78.54, range 35.7-100.00). A notable finding from the Punjab study, all 719 respondents (100%) reported washing their hands with soap and water before and after meals, as well as after defecation. The Universal adherence to hand hygiene is attributed to the behavioural changes and practices adopted during the COVID-19 pandemic. While knowledge of disease transmission remains low, these personal hygiene practices were consistently followed across the surveyed districts (Figure 6). However, there was no significant positive correlation between knowledge and practices scores (r=0.033, p=0.378), indicating the high practice scores were not driven by disease transmission knowledge.

**Figure 6:**
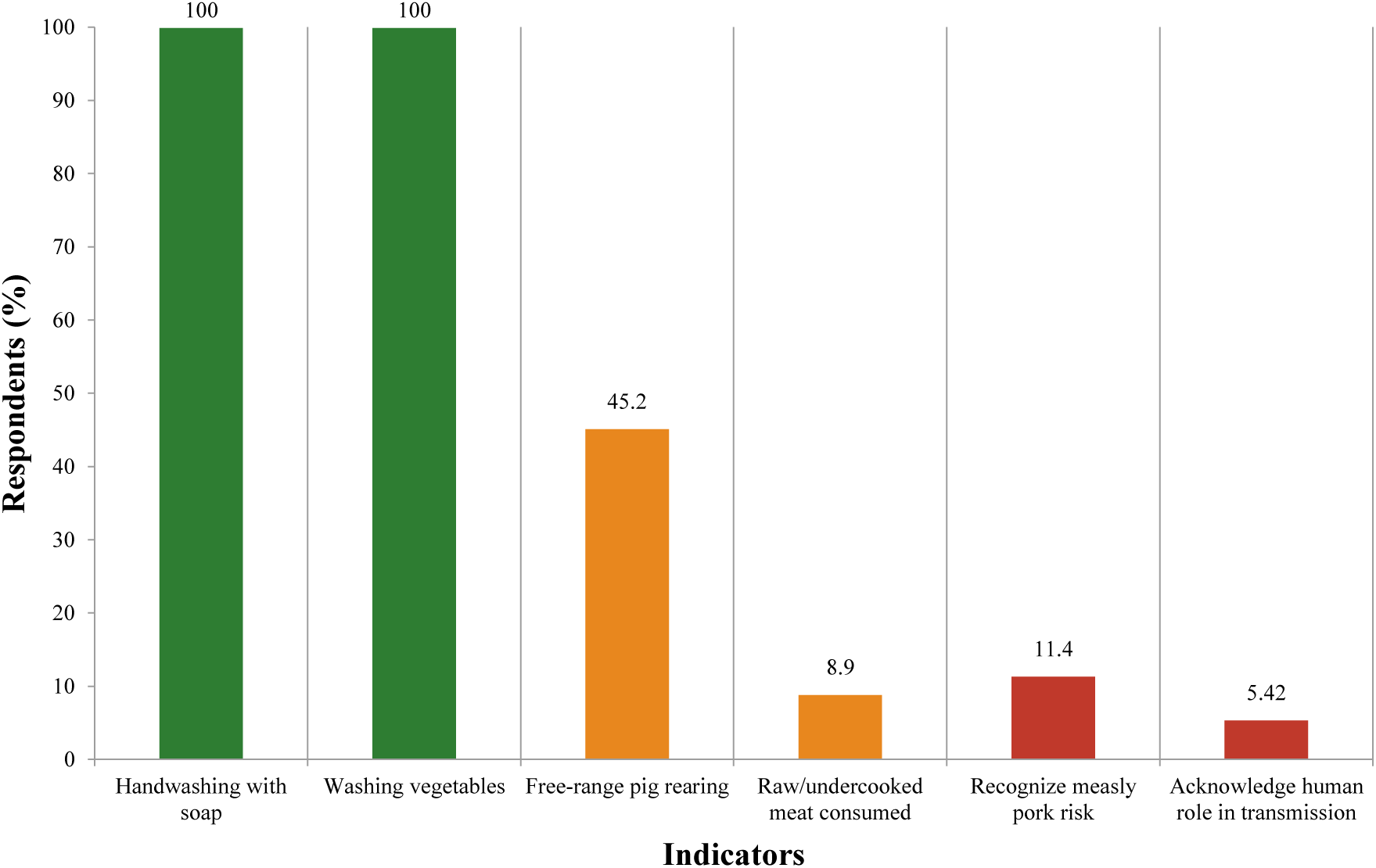
Hand Hygiene practices attributed to COVID-19 behavioural change against persistently risky husbandry practices and critical gaps in meat safety awareness.

### 3.4 Univariate Analysis

The univariate logistic regression revealed pig rearing practices demonstrated a statistically significant association with good knowledge (p=0.010). Intensive farming systems showed 3.46 times higher odds of good knowledge compared to free-range systems (OR=3.363, 95% CI: 0.979-12.261), with descriptive analysis showing 12.50% achieving good knowledge versus 7.43% in free -range and 4.43% in semi-intensive systems. Occupation approached statistical significance (p=0.054) with pig farmers demonstrating 2.73 times higher odds of good knowledge compared to non-farmers (OR=2.730, CI:0.981-7.592), achieving mean knowledge scores of 15.94 ± 8.01 versus 12.49 ±8.69 respectively. Demographic variables including district (p=0.687), gender (p=0.364), age (0.994) and deworming history (p=0.986) showed no statistically significant associations with good knowledge scores (Table 2).

**Table 2:** Univariate Logistic Regression Analysis of factors associated with goof knowledge of *T. solium* infection in Pig farming communities, Punjab.

| S. No. | Variable | Category | Odds Ratio | 95% Confidence interval (CI) | P value |
| --- | --- | --- | --- | --- | --- |
| **1** | Area | Amritsar | - | - | 0.687 |
|  |  | Bathinda | 2.125 | 0.266-22.181 |  |
|  |  | Jalandhar | 1.085 | 0.188-24.008 |  |
|  |  | Ludhiana | 2.547 | 0.111-10.610 |  |
|  |  | Patiala | 2.429 | 0.320-20.240 |  |
| **2** | Gender | Male | - | - | 0.364 |
|  |  | Female | 0.649 | 0.255-1.652 |  |
| **3** | Age | 18-30 | - | - | 0.994 |
|  |  | 31-40 | 0.809 | 0.284-2.307 |  |
|  |  | 41-50 | 1.038 | 0.314-3.438 |  |
|  |  | 51-60 | 0.572 | 0.071-4.608 |  |
|  |  | >61 | 0.000 | 0.000 |  |
| 4 | Education qualification | 0-5<sup>th</sup> | - | - | 0.996 |
|  |  | 6-8<sup>th</sup> | 0.601 | 0.195-1.848 |  |
|  |  | 9-10<sup>th</sup> | 0.745 | 0.166-3.342 |  |
|  |  | 11-15<sup>th</sup> | 0.000 | 0.000 |  |
| 5 | Occupation | Not a Pig farmer | - | - | 0.054 |
|  |  | pig farmer | 2.730 | 0.981-7.592 |  |
| 6 | Deworming | No | - | - | 0.986 |
|  |  | Yes | 1.031 | 0.234-4.539 |  |
| 7 | Pig rearing practices | Free-range | - | - | 0.010 |
|  |  | Semi-intensive | 0.469 | 0.122-1.799 |  |
|  |  | Intensive | 3.464 | 0.979-12.261 |  |
|  |  | Not associated | 0.377 | 0.121-1.167 |  |

Note: OR: Odds ration; CI: Confidence Interval; (-): indicate reference categories used as a baseline for comparison. Add odds ratios are calculated relative to these reference categories; statistical significance was determined using univariate logistic regression analysis with significance level set at $p < 0.05$ .

## 4.0 Discussion

The finding of this study establishes a critical baseline for Taenia solium control within the pig farming communities of Punjab, India. The primary investigation revealed a profound knowledge disparity with only 5.7% understanding the implications of cysticercosis. The lack of knowledge regarding *T. solium* transmission routes mirrors finding in Burkino Faso, where 99.40% of respondents were unaware of how pigs become infested (Dahourou *et al*., 2018). Similarly, in Tanzania awareness of porcine cysticercosis reached 97%, yet understanding of human cysticercosis was virtually non-existent (Nyangi *et al*., 2022; Shonyela *et al*., 2017; Boa *et al*., 2006).

A unique cultural finding in Punjab is the half of the respondents incorrectly attribute cysticercosis to eating raw cabbage rather than pork or human faecal contamination. Nearly half of respondents in a related Indian clinic study believed that eating raw cabbage causes cysticercosis (Sankhyan *et al*., 2015; Girotra *et al*., 2011). A possible explanation is the belief system enforced by unethical local practices where quacks present cabbage worms as proof of human brain infections. Thus, highlighting the need of extensive public health information in form of infotainment or pamphlets that actively deconstructs these specific myths.

A reoccurring theme across the sources is that formal education is a poor indicator of disease knowledge. In Punjab, none of the community members with education above matriculation achieved a good knowledge score, instead direct professional involvement in intensive farming was seen as primary awareness diver. This suggests, further targeted interventions should focus on engagement in the livestock sector particularly pig farming as well selling pork rather than relying on general formal schooling. The aligns with studies in Tanzania and Mexico, where tailored health education packages proved effective regardless of a participant formal education level (Mkupasi *et al*. 2024; Nyangi *et al*., 2025). Interestingly, while many African studies reported that males have higher knowledge scores due to great exposure to husbandry practices (Zulu *et al*., 2023). While study in Punjab suggested that professional proximity to livestock as more critical variable than gender of respondents.

The reported 100% adherence to hand hygiene in Punjab represent a significant outlier compared to other endemic regions. In Zambia, 71% reported handwashing, only 52% actually used soap whereas direct supervision revealed a striking discrepancy between reported toilet use and the actual presence of functional toilet facilities (Zulu *et al*., 2023). However, this shift did not extend ti animal husbandry, as 45.20% of Punjab pig rearer’s continue to use free-range system. The finding is concurrent with observations of Peru and Tanzania, where economic benefits frequently override health knowledge, leading farmers to allow pigs to scavenging despite the risk of porcine cysticercosis (Sankhyan *et al*., 2015).

The level of hygiene adherence in Punjab represents a significant positive shift in public health behaviour, likely driven by government-run awareness programme during the COVID-19 pandemic. The global emphasis on hand hygiene appears to have had a spillover effect, effectively interrupting the faeco-oral transmission cycle not only for COVID-19 but also for *T. solium* and other enteric pathogens.

The sharp contrast in participation interest between Punjab and other regions. While 69.1% of respondents in Pakistan were willing to participants in elimination campaigns of cysticercosis, only 8.8% of participants in Punjab study were personally willing to join a mass drug administration (MDA) programme (Bibi *et al*., 2023; Sankhyan *et al*., 2015). The unwillingness to participate is likely attributed low formal education (60% studied only upto 5^th^ grade), leading to lack of understanding regarding the long-term benefits of preventive chemotherapy.

The study utilized a cross-sectional design, which limits the ability casual relationships or track the long-term evolution of knowledge, attitude, and Practices (KAP). The data rely on self-reported information, which is susceptible to social desirability bias particularly regarding the 100% handwashing rate. Additionally, the lack of species-level identification of parasites in the field and the potential behaviour change of participants when they are being observed, must be considered when interpreting the findings.

## 5.0 Conclusion

This study highlighted the critical awareness gaps and low participation interest in Punjab, necessitates the need of awareness campaigns for improved integrated one health approach. The future policies should prioritize providing village-level slaughter facilities and trained meat inspectors to address the lack of ability of respondents to differentiate between healthy and measly pork (88.60%). The study highlights the need to utilize visual-based education material (like a short film, brochures, pamphlets) to bridge the knowledge gap among farmers with low formal education. Also, leveraging the pandemic-driven momentum in personal hygiene to introduce broader livestock sanitation reforms, implementing community led trust building exercises to overcome resistance to MDA.

## Data Availability

All data produced in the present study are available upon reasonable request to the authors

